# FutureMS Cohort Profile: A Scottish Multi-Centre Inception Cohort Study of Relapsing-Remitting Multiple Sclerosis

**DOI:** 10.1101/2021.04.15.21253274

**Authors:** P.K.A. Kearns, S.J. Martin, J. Chang, R. Meijboom, E.N. York, Y. Chen, C. Weaver, A. Stenson, K. Hafezi, S. Thomson, E. Freyer, L. Murphy, A. Harroud, P. Foley, D. Hunt, M. McLeod, J. O’Riordan, F.J. Carod-Artal, N.J.J. MacDougall, S.E. Baranzini, A.D. Waldman, P. Connick, S. Chandran

## Abstract

Multiple sclerosis (MS) is an immune-mediated, neuroinflammatory disease of the central nervous system and in industrialised countries is the most common cause of progressive neurological disability in working age persons. However, there is significant inter-individual heterogeneity in disease activity and response to treatment. Currently, the ability to predict at diagnosis who will have a benign, intermediate, or aggressive disease course is very limited. There is therefore a need for integrated predictive tools to inform individualised treatment decision-making. FutureMS is a nationally representative, prospective observation cohort study comprising of 440 participants with a new diagnosis of relapsing remitting MS living in Scotland between May 2016 and March 2019. Established with the aim of addressing this need for individualised predictive tools, the cohort is designed to combine detailed clinical phenotyping with imaging, genetic and biomarker metrics of disease activity and progression. Recruitment, baseline assessment and follow up at year one is complete and longer term follow up is planned, beginning at five years after first visit. The study aims to explore the pathobiology and determinants of disease heterogeneity in MS. Here we describe the cohort design and present a profile of the participants at baseline and one year of follow up.

## INTRODUCTION, BACKGROUND AND RATIONALE

Multiple sclerosis (MS) is the leading cause of progressive neurological disability in young and working age persons in middle- and high-income countries, and is a paradigm of neuroinflammation, autoimmunity and neurodegeneration^1^. 85-90% of incident cases have a relapsing-remitting disease course (RRMS) at onset, characterised by periods of clinical symptoms emerging and resolving. After a median of approximately 20-years, the disease moves into a phase of progressively accumulating irreversible disability called secondary progressive MS. The other 10-15% of incident cases experience this progressive phase from onset (primary progressive MS). In the RRMS group, both inflammation and neuronal injury are present throughout the disease course, with multifocal inflammatory demyelination dominant in the RR phase and neurodegeneration the key pathological substrate of the progressive phase^1^. Whilst the disease remains incurable, and untreated typically results in accumulation of substantial disability and a 5-15 year reduction in life expectancy, the emergence of effective disease modifying therapies (DMTs) for the early phase of disease has transformed the outlook for people living with RRMS in recent years^2,3^.

rMS has a markedly heterogenous natural history; cases of aggressive progression and relatively indolent disease occur on a spectrum even in untreated individuals. The ability to predict future disease activity for an individual is very limited and still reactive in practice, extrapolating from radiological or clinical evidence of past disease activity. Informed treatment and lifestyle decision-making by people newly diagnosed with MS requires predictive tools available at or close to the point of diagnosis. Increasing DMT options, with attendant side effect profiles, highlights the urgent need for accurate and personalised prognostic tools.

DMTs do not appear to treat all biological aspects of MS pathology equally. They are more effective at preventing neuroinflammation than at halting neurodegeneration. Neurodegeneration is also hard to measure over short time periods, complicating early prediction. It may be essential for personalised predictive tools to be capable of discriminating between different biological contributions to disability progression. Early predictors and determinants of neuroinflammation may differ from those predicting the rate of neurodegenerative disease activity. In order to unpick this complexity, long-term longitudinal follow up of adequately-powered and representative clinical cohorts, starting as early as possible in disease course, which are resourced to “deeply phenotype” participants, could be a vital contribution towards achieving this personalised decision making.

One such cohort, FutureMS, is now fully recruited in Scotland and the first follow-up wave at one year has been completed. FutureMS is a large (n = 440) prospective inception cohort study recruiting newly diagnosed persons with relapsing-remitting MS (RRMS) living in Scotland at the time of their diagnosis. With a high incidence of MS, a stable population of 5.4. million, low rates of migration, and a national single-payer universal healthcare system free at the point of use, Scotland offers an ideal setting for a long-term longitudinal study of pwMS^4^.

The FutureMS study hypothesis is that inter-individual variability in disease activity in RRMS is determined and will be predictable by a combination of clinical, laboratory, imaging and genetic parameters. The primary aim is to develop predictive tools for focal neuroinflammatory disease activity based on clinical, laboratory, magnetic resonanceimaging (MRI) and genomic assessment in patients with RRMS. Secondary outcomes include the development of predictive tools for a) neurodegenerative disease activity, b) clinical measure of disease activity, and c) clinical measures of quality of life. The study is structured in waves at baseline (within six months of diagnosis), at month 12 (baseline + 12 months), and future follow-up is planned at five years (baseline +5 years), and subsequently thereafter. Given that MS is a chronic long-term condition, it is expected that further insights will emerge from study of the cohort over time. Future MS aims to reduce uncertainty in disease trajectory and to allow for more tailored and personalized care for pwMS. This paper is intended to provide an overview of the study design and introduce a profile of the study participants.

### PARTICIPANTS Representative cohort

Between May 2016 and March 2019, 440 adult patients (age ≥ 18 years) were recruited as a nationally representative incidence sample within six months of their diagnosis (median time since diagnosis at first study visit: 60 days, IQR: 61 days). To ensure national representativeness, the study was designed to support inclusion of any person newly diagnosed with RRMS wherever they may live in Scotland, aiming to establish both geographically and socioeconomically representative coverage of the Scottish mainland and islands.

Participants were recruited from the five main tertiary Scottish clinical neurology centres: 185 from Edinburgh (42.0%), 164 from Glasgow (37.3%), 46 from Dundee (10.5%), 35 from Aberdeen (8.0%), and 8 from Inverness (1.8%). This roughly reflects the geographic distribution of the population of Scotland, and the geographic incidence burden of MS^4^ (see. Fig 1.). Analysis of the Scottish Multiple Sclerosis Register (SMSR)), a national incidence register with mandatory-reporting for MS specialist nurses at the time of referral for newly diagnosed persons, reveals that 45% of all persons diagnosed with RRMS in Scotland over this period were recruited to FutureMS. Comparison with the demographic characteristics from the SMSR suggests a broadly representative sample was recruited (table 1). FutureMS participants were slightly younger on average, less represented at the extremes of age distribution (Fig S1.) and more likely to be female. As has been observed in the SMSR data, there was a significant excess of persons living in affluent Scottish Index of Multiple Deprivation^5^ (SIMD) quintiles relative to deprived quintiles (SIMD: X^2^ = 14.06, 4d.f., p<0.01; and FutureMS: X^2^ = 12.2, 4 d.f., p <0.05) (Fig S2). This has been recognized in multiple epidemiological studies in Scotland^6,7^ and is not apparently explained -- in fact paradoxical -- given the burden of established MS environmental risk factors (e.g. Vitamin D deficiency, obesity in adolescence and smoking), which are strongly all associated with deprivation in Scotland^8–11^.

**Table 1.** Comparison of the baseline demographics. Data for persons with RRMS recorded in between Jan 1^st^ 2010 and Dec 31^st^ 2017 Scottish MS Register^4^. P-values for test hypothesis that there is no difference in proportions or means, between the two study populations, calculated by Chi-squared test (proportions) or two-sided t-test (age).

|  | FutureMS | Scottish Multiple Sclerosis Register | p-value |
| --- | --- | --- | --- |
| **Female (n, %)** | 325 (73.9%) | 1916 (71.3%) |  |
| **Male (n, %)** | 115 (26.1%) | 772 (28.7%) | .26 |
| **SIMD Quintile** |  |  |  |
| **1 (most deprived)** | 71 (16.1%) | 671 (18.2%) |  |
| **2** | 75 (17.0%) | 718 (19.5%) |  |
| **3** | 84 (19.1%) | 719 (19.5%) |  |
| **4** | 102 (23.2%) | 802 (21.8%) |  |
| **5 (least deprived)** | 108 (24.5%) | 770 (20.9%) | .29 |
| **Age at symptom onset (mean (range))** | 33.8 (50.69) | Data not available |  |
| **Age at diagnosis (mean (range))** | 37.7 (48.3) | 38.1 (64.8) | .49 |

**Figure 1:**
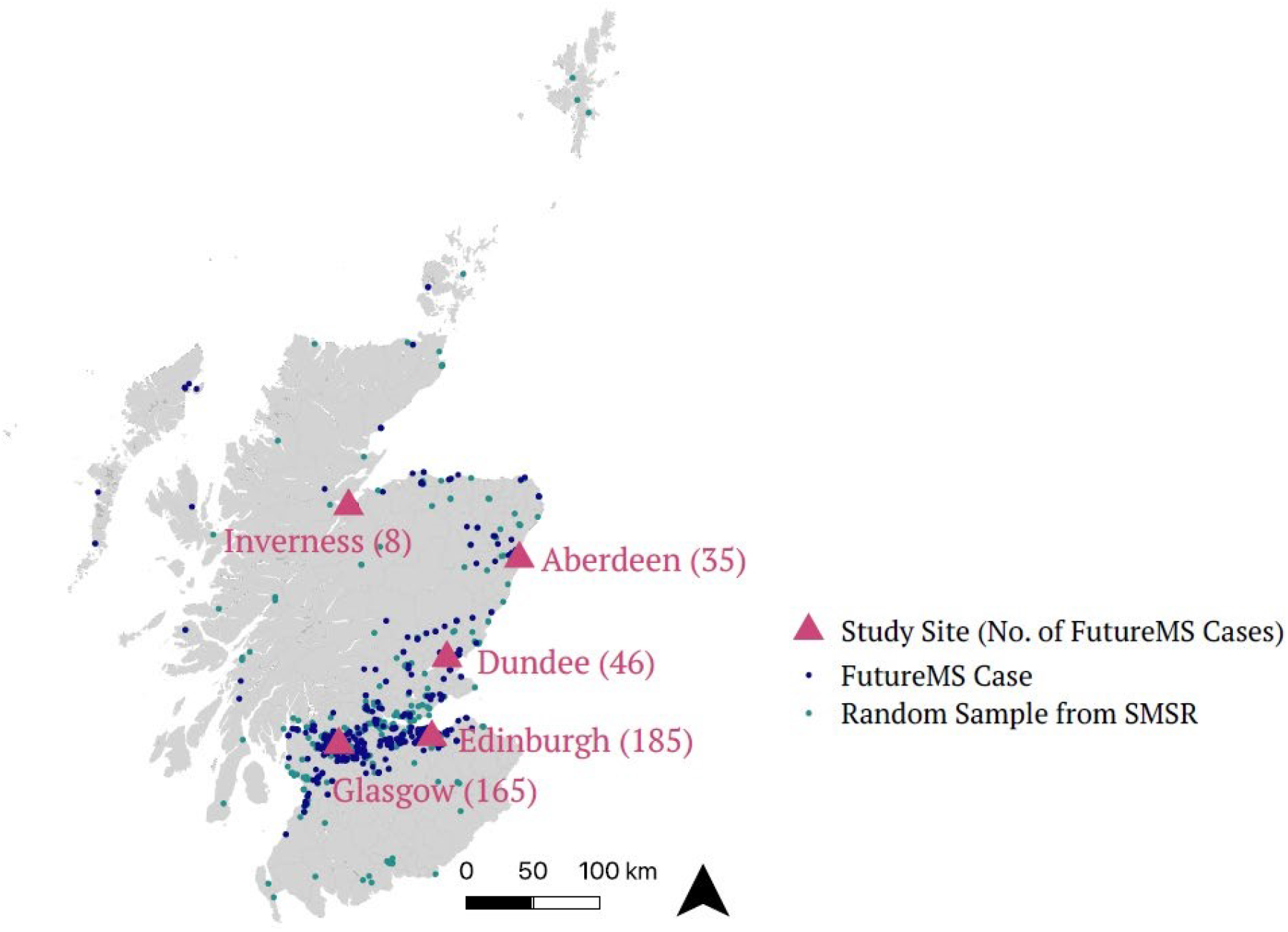
Map of FutureMS participants by approximate location of residence at the time of diagnosis. Participant locations are not precise, located at the population centroid of the nearest SIMD intermediate zone (mean population ∼4000). FutureMS cases (purple) are displayed alongside intermediate zone of residence of a random selection of 440 individuals from the SMSR (green). All map positions have latitudinal and longitudinal random noise added to prevent personal identifiability.

Amongst FutureMS participants who listed their ethnicity, 426/440 (96.8%) recorded their primary ethnicity as White Scottish/British. For those with recorded ethnicity in the first eight years of the SMSR (2010-2018), the proportion recorded as Scottish, British or Irish was similar 862/919 (93.8%).

### Diagnostic inclusion criteria

Diagnosis in all cases was confirmed by the treating consultant neurologist as fulfilling the most recent McDonald Criteria. Participants must not have commenced on DMT prior to baseline assessment, had capacity to give informed consent and have had no contraindication to MR brain imaging at the time of their baseline visit.

Only patients with a diagnosis of RRMS were eligible for inclusion in FutureMS. We excluded those with progression at onset. Diagnosis of progressive forms of MS is typically delayed relative to RRMS, requiring a period of observation of sustained progression, and recent studies have suggested that the distinction between relapsing and progressive forms of MS are not clearly demarcated clinical entities. Rather that they reflect different stages of the same disease^12^., inclusion was limited to persons with RRMS.

## CONTROLS FOR LABORATORY AND BIOMARKER STUDIES

103 healthy volunteers were recruited from the Lothian area to donate blood and DNA for the biomarker and genetic analyses. These persons were age- and sex-frequency matched to the study population. All were recruited in the Anne Rowling Regenerative Neurology Clinic in Edinburgh, and so are mainly drawn from the surrounding Edinburgh and Lothian areas.

## STUDY VISITS

Study visits were in addition to, and not in place of, standard neurological care. Consequently, the timing of the first visit is likely to be associated with temporal fluctuations in the participants’ disease activity, as diagnosis is more likely following clinical relapse. Subsequent visits should be independent of this potential bias being at fixed intervals after first visit and not triggered by clinical disease activity. We consider it a strength that the study design reduces clinically-triggered follow up biases.

All clinical management decisions were reserved to the treating team. Participation in FutureMS is not a barrier to participating in any other research study including interventional trials, and we anticipate that a substantial number of participants will choose to engage with other research studies. **[Figure 2 approximately here]**

**Figure 2.**
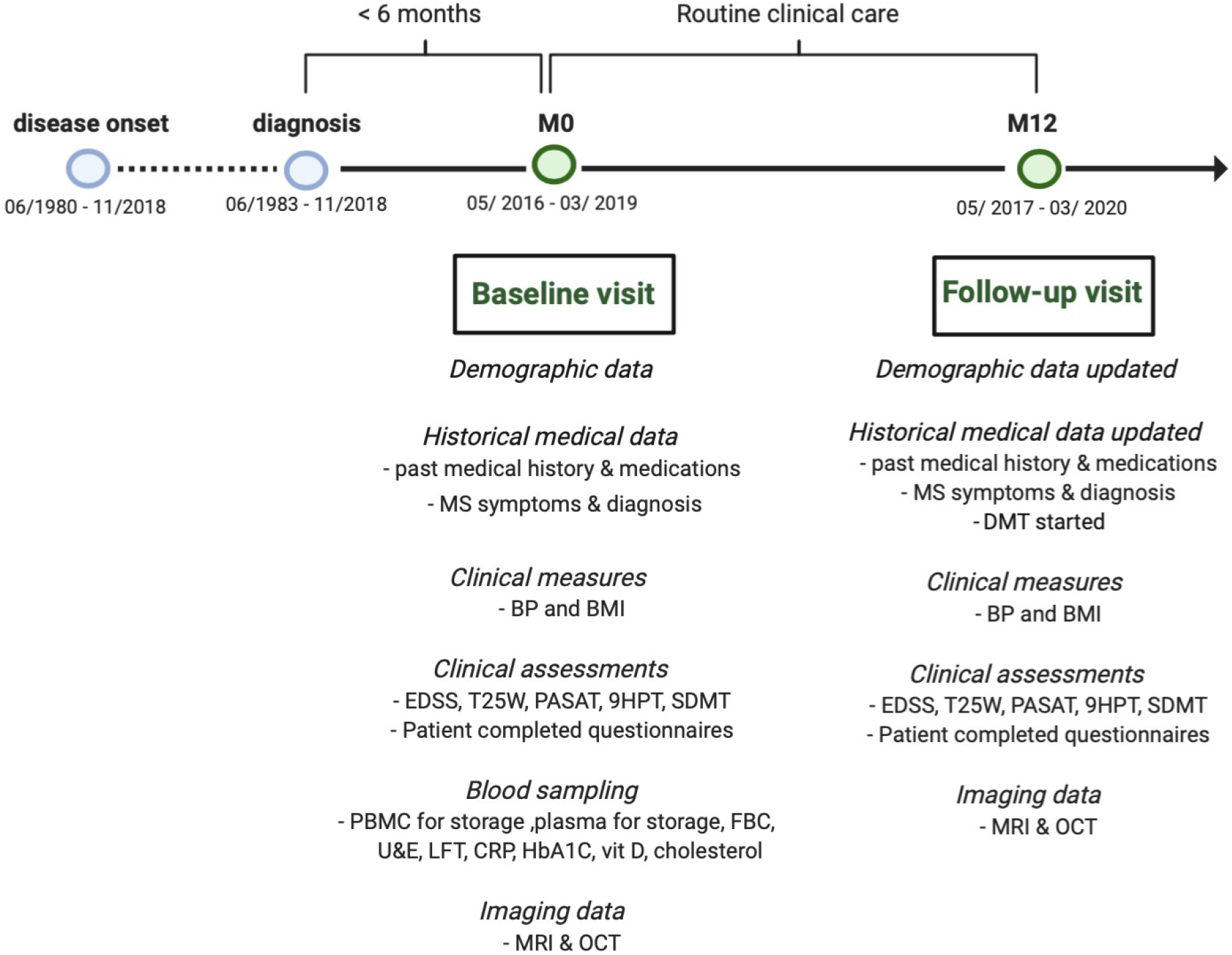
FutureMS cohort design. BP – blood pressure; BMI – Body mass index; EDSS – Extended disability status scale; T25W – Timed 25foot walk test; PASAT – paced auditory serial addition test 3; 9HPT – nine hole peg test; SDMT – symbol digit modality test; PBMC – Peripheral blood mononuclear cells; FBC – full blood count; U&E – urea and electrolytes and renal function tests; LFT – liver function test panel; CRP – C-Reactive protein; HbA1C – glycosylated haemoglobin A1C; vit D – 25-OH-vitamin D; MRI – magnetic resonance imaging; OCT – optical coherence tomography.

Demographic and clinical variables collected at baseline included date of birth, sex, ethnicity, occupation, co-morbidities, medication history (including ‘over the counter’ and supplements), and family history. Data pertaining to the diagnosis of RRMS (description of initial symptoms, number of clinical relapses, hospitalisations, and steroid use) were recorded at baseline visit, and all data were also updated at the twelve-month review (Table S1).

At the time of writing, 392 of 440 participants (89.1%) had completed the return visit at year one. Six patients had withdrawn consent (1.4%), 23 (5.2%) were lost to follow up but will be invited to participant in subsequent visits. 19 (4.3%) participants had their return visit at one year prevented by the COVID-19 pandemic (visits due in March or April 2020) but these participants will be invited for future follow up.

### Sub-studies

Alongside the main study, four additional ‘opt in’ sub-studies allowed deeper phenotyping of participants. Sub-studies included consenting participants to approach them with opportunities for future research/cross-linkage with other studies (sub-study 1); biobanking an additional large volume blood sample at baseline visit (sub-study 2); retinal imaging with optical coherence tomography at baseline and twelve-month follow-up visit (sub-study 3); and additional advanced MRI imaging sequences (sub-study 4, baseline n = 78, follow up n = 74, complete pairs n = 67).

## CLINICAL OBSERVATIONS

Patient-reported and assessor-measured clinical observations were collected at each study visit. Source data from clinical assessments at local sites was captured using a web-based electronic case report form (eCRF). Both participants and study staff entered data directly. Clinical data were entered by participants via questionnaires. There was a high level of engagement with these questionnaires and assessments. Data completeness was >99% across all clinical measured and reported variables at both the baseline and year 1 visits.

Questionnaires included the multiple sclerosis impact scale (MSIS-29)^13^, NICE domain activity & impairment, CDC Health Related Quality of Life (HRQOL-4)^14^, Patient Determined Disease Steps (PDDS)^15^, Fatigue Severity Scale (FSS)^16^, Generalized Anxiety Disorder Assessment (GAD-7)^17^, depression assessment scoring (PHQ-9)^18^, Baecke Habitual Physical activity, cognitive, leisure, social and lifestyle questionnaires. Clinical measures included the Expanded Disability Severity Score (EDSS)^19^, components of the Multiple Sclerosis Functional Composite score (MSFC)^20^, mean arterial blood pressure (BP) and Body Mass Index (BMI).

On some measures participants improve on average over the first year. This is not unexpected as diagnosis often coincides with a clinical event/relapse and we expect some regression toward the mean over the course of the first year which may be amplified by effective treatment for some participants. Many of these clinical measures and questionnaires capture overlapping phenomena and are correlated. This provides both opportunities and challenges for asking causal questions of longitudinal repeated measures (see Fig S4).

## RESULTS

### Lifestyle and Social Factors

Lifestyle and social factors are known to influence MS disease course, and whilst some of these factors have been identified, there is much non-heritable variability that remains unexplained. Amongst the strongest known environmental factor is smoking^21^. There is strong evidence that smoking both modifies the risk of MS incidence and affects the rate of disability progression. Smoking has also been demonstrated to interact statistically with disease risk loci^22^. In our cohort, 14.7% of participants were current smokers at the baseline visit and more than half (50.7%) declared themselves to be “ever smokers”. By year one, there has only been a modest reduction in participants smoking (13%). Figure 3A demonstrates that even at baseline, the distribution of disability differs by smoking status.

**Figure 3:**
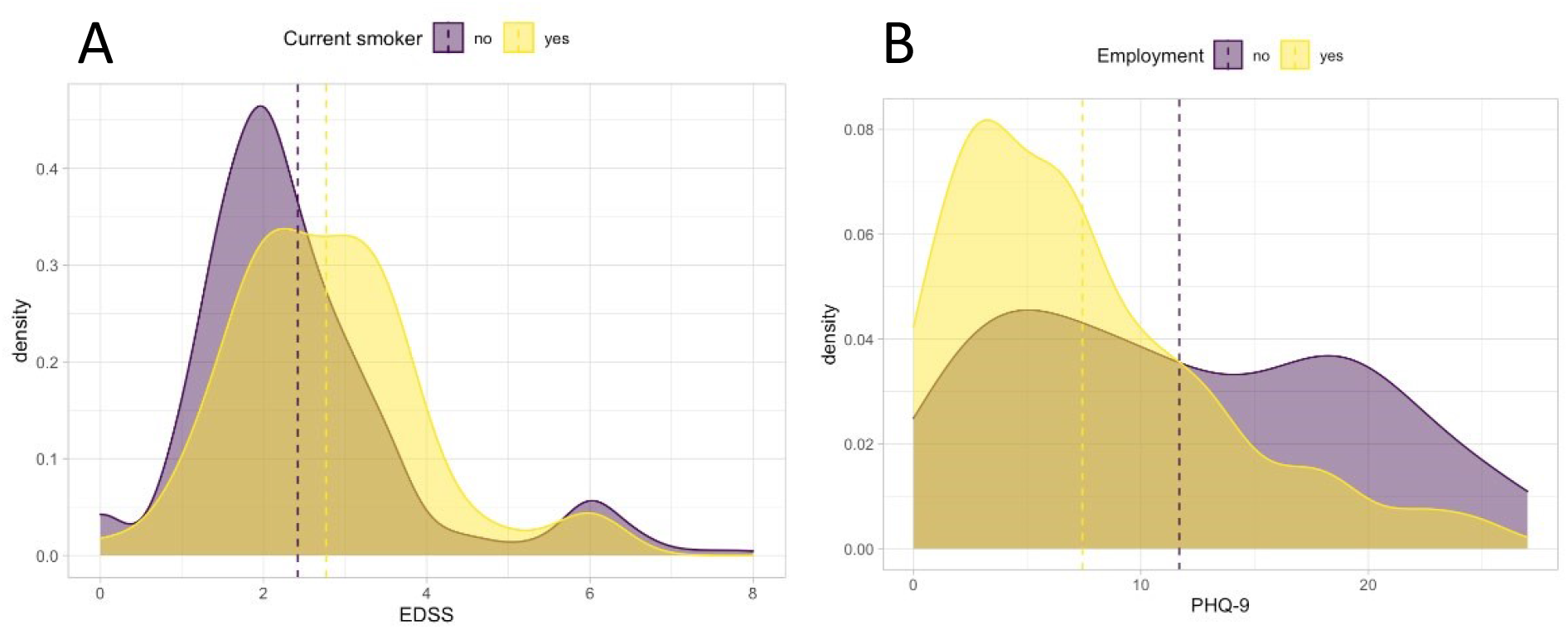
Density plots stratifying the cohort at baseline visit. A – Distribution of EDSS (a measure of physical disability) by smoking status. B – Evidence of greater burden of depression as detected by PHQ9 in those who are unemployed at baseline.

As MS is associated with significant disability in working-age persons, it has the potential to impact on employment and the quality of working life. Figure 3B demonstrates that at baseline the distribution of depression scores (PHQ-9) for persons who are employed and unemployed is quite different. The bimodal distribution of those who are unemployed, suggests that a significant proportion of pwMS who are unemployed at baseline are at risk of depression as assessed by this score.

### Physical disability

The distribution of measures of physical disability follow similar patterns across the two waves of the study cohort and there is little difference between these over this early period. This demonstrates the relatively insensitive nature of these measures early in disease course and over shorter study periods, at least at population level. However, FutureMS is sufficiently powered to allow meaningful comparison of sub-cohorts and of outlier individuals whose measured scores have worsened or improved in the first year. Figure 5 demonstrates one such analysis: the group who have worsened over the first year appear older and very few have low fatigue severity scores at baseline. Further analyses of these patterns may define groups that explain some of the heterogeneity in disease course.

**Figure 4:**
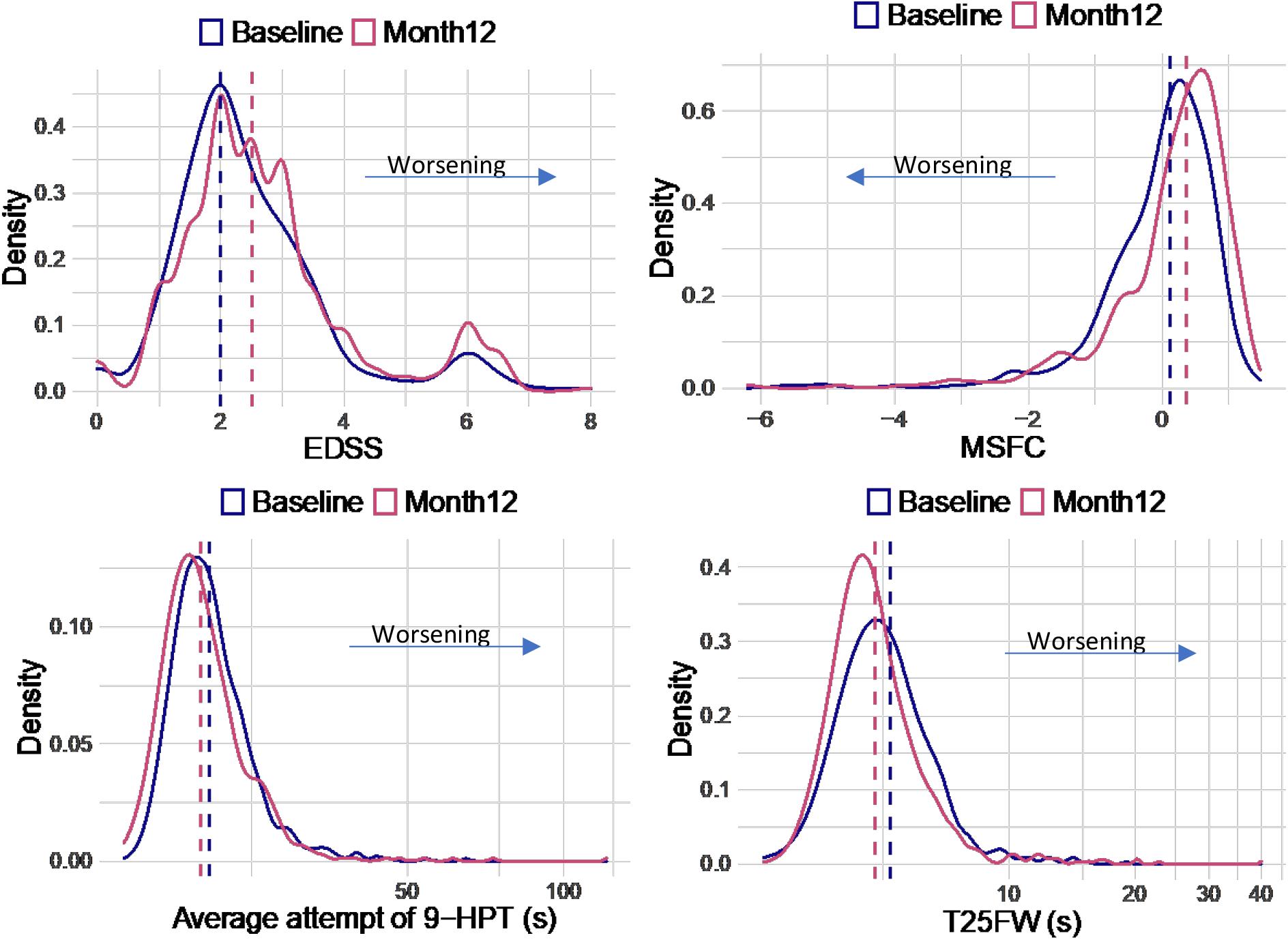
Physical measures of disability across the cohort at baseline and month 12. T25FtWT X axis is in seconds, 9-HPT is the mean between hands of the mean of two attempts with each hand and is a measure of upper limb disability measured in seconds (longer time reflects less dexterity). EDSS is an ordinal scale where higher scores reflect greater disability. MSFC is a continuous scale(z-score), participants who are unable to walk are arbitrarily attributed very low Z-scores for the walking component of their test (−13.7) as per published instructions. This gives a long negative tail to the distribution as the -13.7 is chosen to allow for the cohort to progress in disability with time and still capture variance in walking ability. However, this is evidently somewhat distortionary.

**Figure 5:**
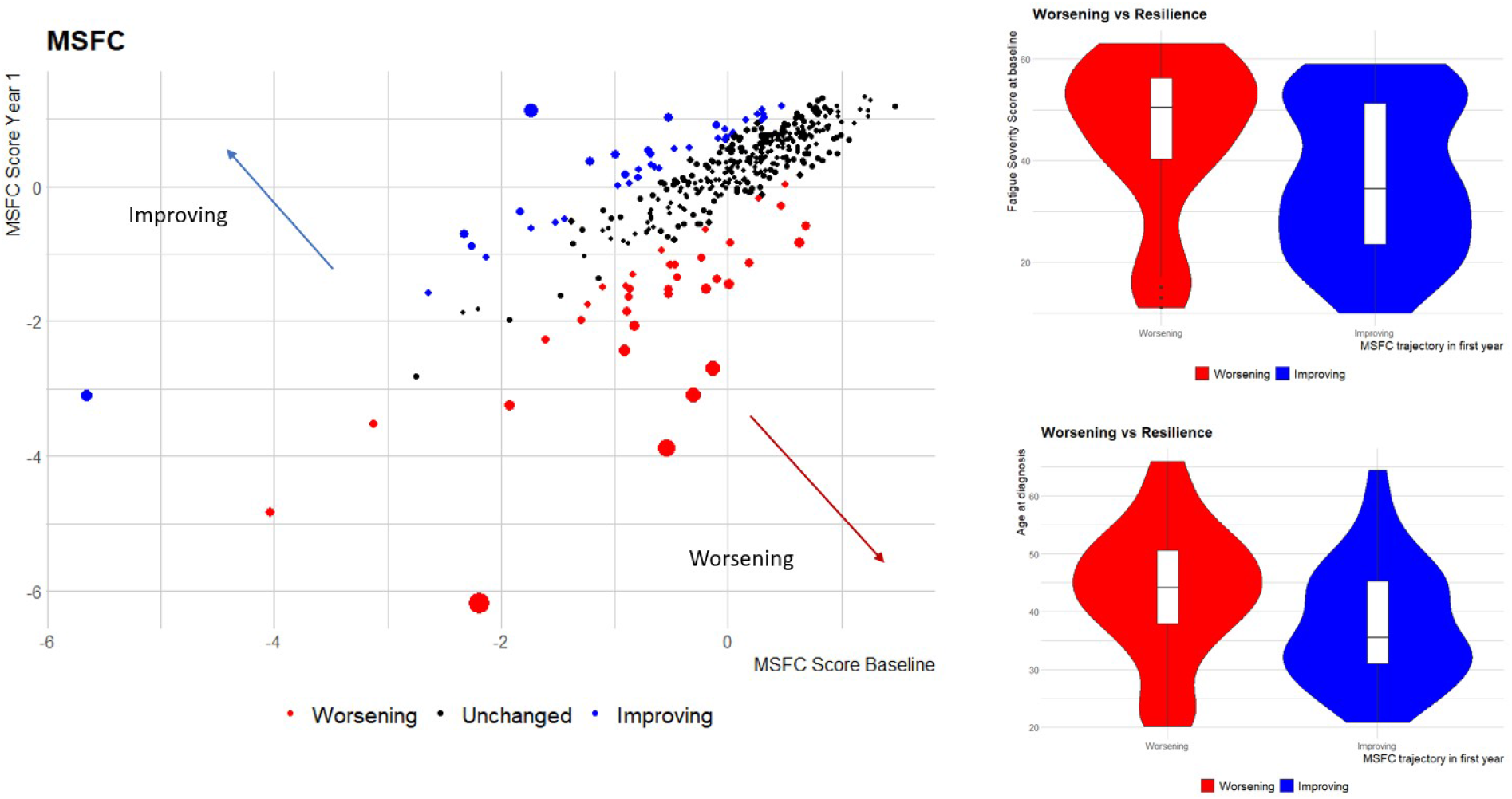
Individual level change in physical disability between the waves. Circle size reflects size of difference between MSFC measurements between study visits (squared residual from least squares regression line of MSFC at year one on MSFC at year two). Outlier groups defined as above the 90^th^ and below the 10^th^ centile for regression residual.

### Fatigue, cognition and mood

Fatigue has been described as the most disabling MS symptom by as many as 60% of patients in some studies^23^. The fatigue severity scale is therefore an important component of the study assessment of MS disease impact. The biological basis of fatigue is poorly understood. The distribution of participants suffering fatigue changes between the baseline and month 12 in our cohort. At follow up, the group does not appear to be monomodal which may reflect underlying biological heterogeneity or discrete sub-populations. Previous work has attempted to stratify fatigue into central or peripheral fatigue, and it may be that fatigue is a composite symptom with multiple pathogenic mechanisms. Investigation of the natural history, burden and biology of fatigue will be a focus of study in the FutureMS cohort.

We observed a significant burden of depression in the study cohort, as measured by PHQ-9, highlighting the important contribution of mental health to MS burden early in the natural history of the MS. Median depression scores improved statistically significantly over the course of the first year from 7 to 4 (p<10^−12^, two-tailed Wilcoxon signed-rank test).

Cognitive impairment measured by SDMT and PASAT-3 tests revealed marked heterogeneity in the burden of impairment and heterogeneity in the trajectory of these measures between study waves. Whilst the SDMT and PASAT-3 scores are significantly correlated (Spearman’s rho 44.7 at baseline and 48.7 at follow up, both p<10^−15^), we found some participants struggled with PASAT-3 with those who fail the trial run recorded as zero contributing to a distribution of scores that was far from normal. Cognitive scores at baseline and follow up correlated statistically significantly (p<10^−15^) for both tests: SDMT (rho = 0.8) and PASAT-3 (rho = 0.76).

### Adjusted measures of MS Severity

The multiple sclerosis severity score (MSSS) and age-related multiple sclerosis severity scale (ARMSS) have been validated on large independent cohorts to attempt to standardize physical MS-related disability for disease duration. The MSSS does this by normalising the EDSS for patient-reported disease duration. However, factors that influence recall of disease duration and EDSS may confound this measure. Therefore, ARMSS normalises EDSS for patient age which is correlated with disease duration imperfectly but is not susceptible to recall biases. These measures, ARMSS and MSSS, correlate strongly and statistically significantly (all comparisons p<10^−15^) in both study waves: rho = 0.69 at baseline and rho = 0.71 at year one (figure 7). The age-adjusted measures show good overall agreement with the participant-reported measure (MSSS).

**Figure 6.**
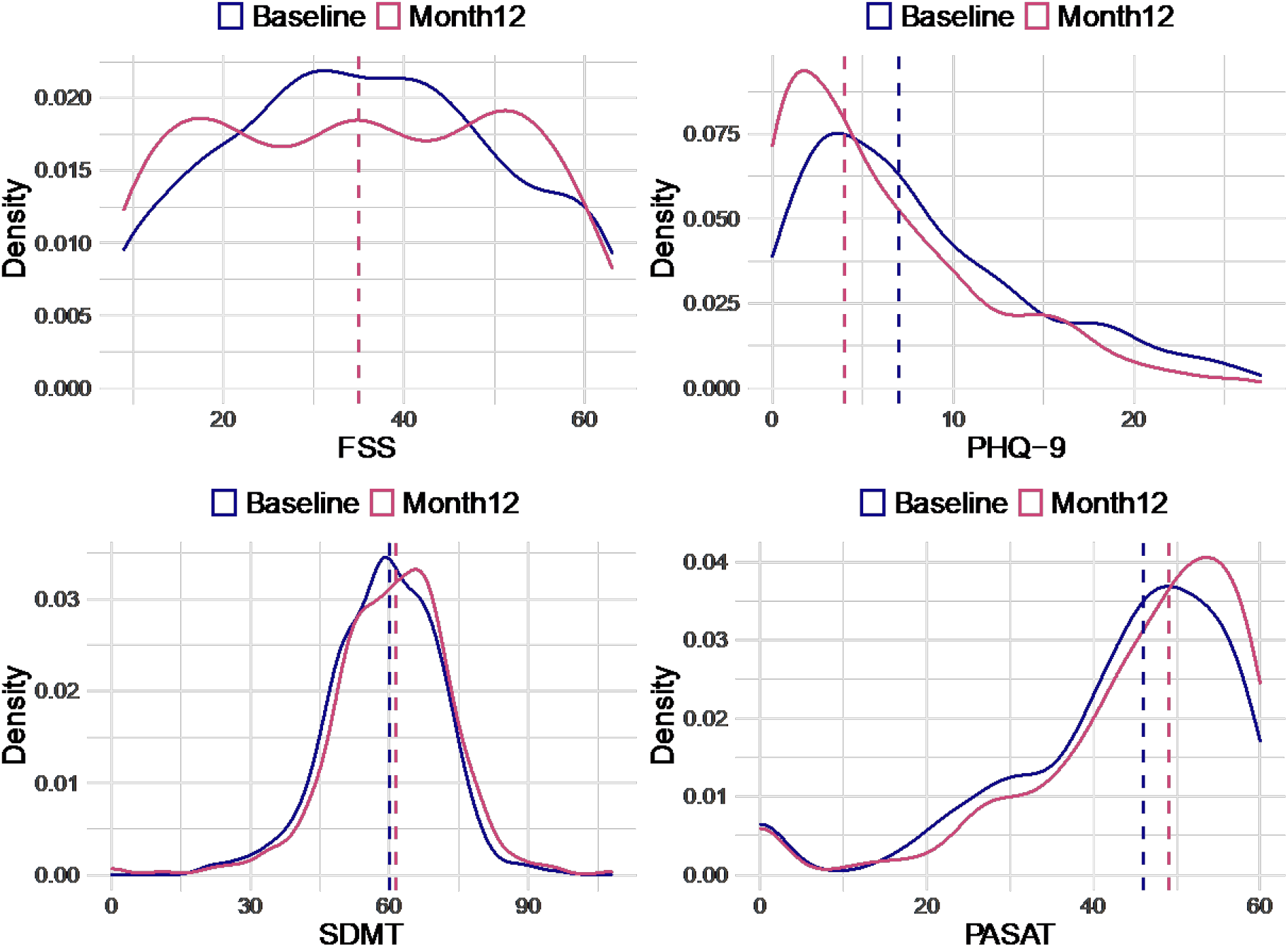
Fatigue Severity Score, PHQ-9 screening tool for depression, Symbol Digit Modality Tool, Paced Serial Addition Tool. Higher scores on Fatigue Severity Scale indicate worse fatigue. Higher scores on PHQ-9 indicate risk of depression. Higher scores on PASAT and SDMT indicate better performance on cognition testing and so less impairment.

**Figure 7.**
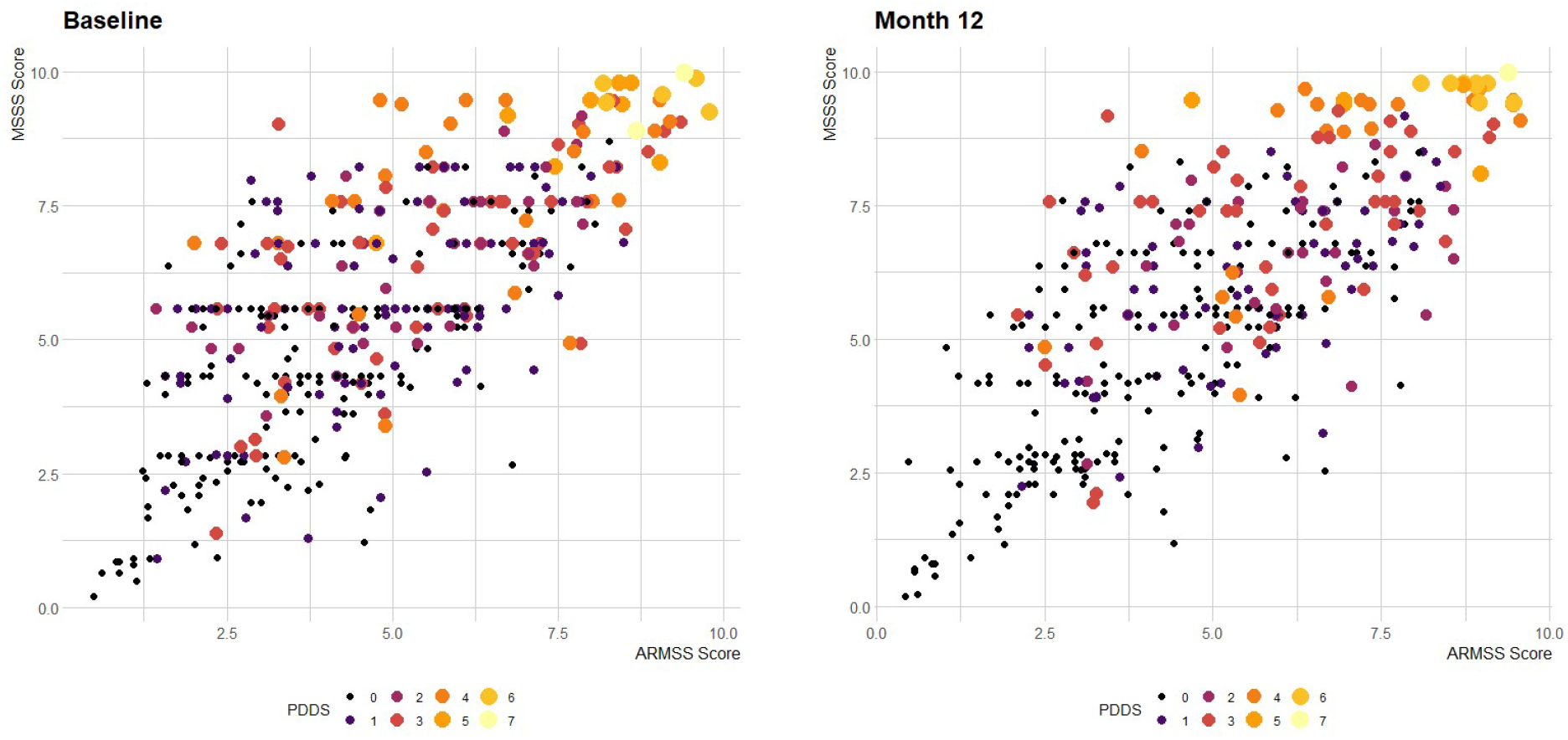
size and colour of the points reflects the patient determined diseases steps. In these figures, points are study individuals, and the size and colour of the points are scaled using the PDDS (range 0 – 7, where 7 is most severe).

## MR IMAGING

MR brain image protocols and processing have been described in detail elsewhere [Meijboom et al, pre-print.], but in brief, participants from all centres were invited to undergo a standard protocol of structural 3T MRI sequences, including T1-weighted, T2-weighted and fluid attenuated inversion recovery (FLAIR) images. The study was powered to detect changes in brain imaging outcomes - not necessarily changes in clinical measures - at year one, as MR brain imaging measures have higher sensitivity over short time frames compared to clinical measures (e.g. annual number of relapses). The primary endpoint was new and/or enlarging T2 hyperintense lesions, as qualitatively (visually) assessed by expert neuroradiologists using brain imaging software. The secondary endpoint was automated measurement of global brain volume change. In addition to the standard structural sequences, participants in Edinburgh were invited to undergo an advanced MR imaging protocol, comprising diffusion MRI (dMRI) and magnetisation transfer imaging (MTI). These measures allow for quantitative assessment of brain microstructure and therefore provide the opportunity to study brain myelin and axonal damage, which are prominent features of MS. dMRI and MTI metrics were used as exploratory endpoints of microstructural change in MS.

## OPTICAL COHERENCE TOMOGRAPHY

Participants of FutureMS were offered the opportunity to enrol in a sub-study of retinal imaging and optical coherence tomography. Proof of concept has been established in MS for the utility of retinal imaging with optical coherence tomography (OCT) to measure thinning of the retinal nerve fibre layer (RNFL), inner nuclear layer (INL), and the ganglion cell and inner plexiform layer (GCIP), all of which have been shown to correlate with clinical activity and disability^24^.

## LABORATORY INVESTIGATIONS

Blood sampling was performed at the baseline visit for routine laboratory testing, genetic testing, cell subsets, and biobanking for future studies. ‘Routine’ analysis included eGFR, HbA1c, CRP, vitamin D, albumin, cholesterol, high-density lipoprotein (HDL), low-density lipoprotein (LDL), very-low density lipoprotein (VLDL), haemoglobin concentration (Hb), white cell count (WCC), and platelet count. All clinical and laboratory (blood test) assessments were performed in a standard sequence by the assessing neurologist or clinical research nurse. All samples were transported to laboratories for analysis immediately after venepuncture. Routine laboratory predictors were analysed in local NHS labs using their standard local protocols (Fig S3.).

Blood was collected at baseline for DNA extraction and PBMC isolation. DNA was extracted from 9ml EDTA whole blood using Nucleon BACC3 kit. DNA samples were re-suspended in 1ml TE buffer pH 7.5 (10mM Tris-Cl pH 7.5, 1mM EDTA pH 8.0). Peripheral blood mononuclear cells (PBMC) were isolated from Lithium Heparin blood at each hub and samples shipped to the Edinburgh CRF Genetics Core Laboratory for storage.

For fluorescence activated cell sorting (FACS), samples and sorted PBMC populations were kept on ice at all times. Prior to sorting on the BD FACSAriaII SORP cell sorter, the instrument was set-up using the internal Cytometer Set-Up and Tracking (CS&T) system, the drop delay was set to >99.9% with Accudrop beads to ensure sort quality. The Aria was set up with the 85um nozzle and 45psi pressure. Single stained controls were analysed with every run before a sort and compensation adjusted if necessary. The sample chamber and collection tube holder were cooled to 4°C. Collection tubes were pre-coated with 500uL cold medium. According to protocol, 5uL of 7-AAD were added to the cell sample 5 minutes before sorting, and the samples were filtered through 35um nylon mesh cell-strainers to avoid the risk of clumping samples interfering with the sort. Gates were set on FSC-H and FSC-A to determine single cells, SSC-A and FSCA to exclude debris and 7-AAD negative population to exclude dead cells 7-AAD+. The populations sorted were CD3+ CD4+ T-cells, CD3+ CD8+ T-cells, CD14+ Monocytes and CD19+ B-cells. The cells were run with a flowrate of 6.000 – 8.000 events per second. The maximum number of cells sorted per population was set to 1.5 × 10^6^ for the larger populations, and as many as possible for the smaller populations. Upon completion of the sort, a different sorted population from each sample was reanalysed on the instrument to evaluate the post-sort purity of the fractions across the samples. The number of cells sorted were recorded and sorted populations passed for RNA extraction.

The fluorescent channels used were: 7-AAD excitation laser 488nm, 685/35nm BP filter, CD3-APC excitation laser 640nm, 670/14nm BP filter, CD14-FITC excitation laser 488nm, 525/50nm BP filter, CD19-BV excitation laser 405nm, 450/50nm BP filter, CD4-PE excitation laser 561nm, 582/15nm BP filter and CD8-BUV excitation laser 355nm, 450/50nm BP filter. RNA was extracted from sorted cell fractions using Qiagen miRNeasy. Yield and RIN were measured by Qubit RNA HS and Agilent Fragment Analyser. 1ng of each total RNA sample was fragmented and first-strand cDNA was generated using the SMARTer Stranded Total RNA-Seq Kit - Pico Input Mammalian kitIllumina-compatible adapters and indexes were added via 5 cycles of PCR. AMPure XP beads (Beckman Coulter) were used to purify the cDNA library followed by ribosomal RNA depletion using ZapR and R-Probes. Uncleaved fragments were enriched by 15 cycles of PCR before a final library purification using AMPure XP beads and sequencing on an Illumina NovaSeq.

Summary results for cell proportions suggested significant inter-individual variability in the proportion of viable cells that were B cells, monocytes, or T cells (supplementary materials) despite the lack of DMTs in this cohort. Future work is intended to explore whether these are meaningful parameters predictive of current or future neuroinflammatory activity.

Additional (fluid) biomarkers of neuroinflammation have been analysed at baseline and will be described in detail elsewhere. These include neurofilament light chains (NfL), GFAP, Tau, UCH-L1 measured using digital ELISA/Single Molecule Array (SIMOA). CSF biomarkers have also been analysed for a subset of study participants.

## SNP GENOTYPING

Although environmental factors (particularly EBV infection, smoking, obesity during adolescence) are known to make important contributions to MS risk, there is an important heritable component evidenced from correlation between relatives^25^. The strongest known contribution to this heritability is for the HLA region of chromosome 6^26^. Despite the remarkable allelic heterogeneity observed at this region, HLADRB1*15:01 (marked by rs3135388^27^ and several SNPs in high linkage disequilibrium) is known to dominate the contribution to this risk. However, in addition, over 200 non-HLA loci are associated with disease risk^28^. Less is known about genetic contributions to the variance of disease course. Figure S5 demonstrates the high confidence in calling (discriminating between) SNP genotypes linked to HLADRB1*15:01 in FutureMS and table 3 demonstrates the expected finding of significant overrepresentation of the HLA DRB1*15:01 risk loci.

**Table 2.**
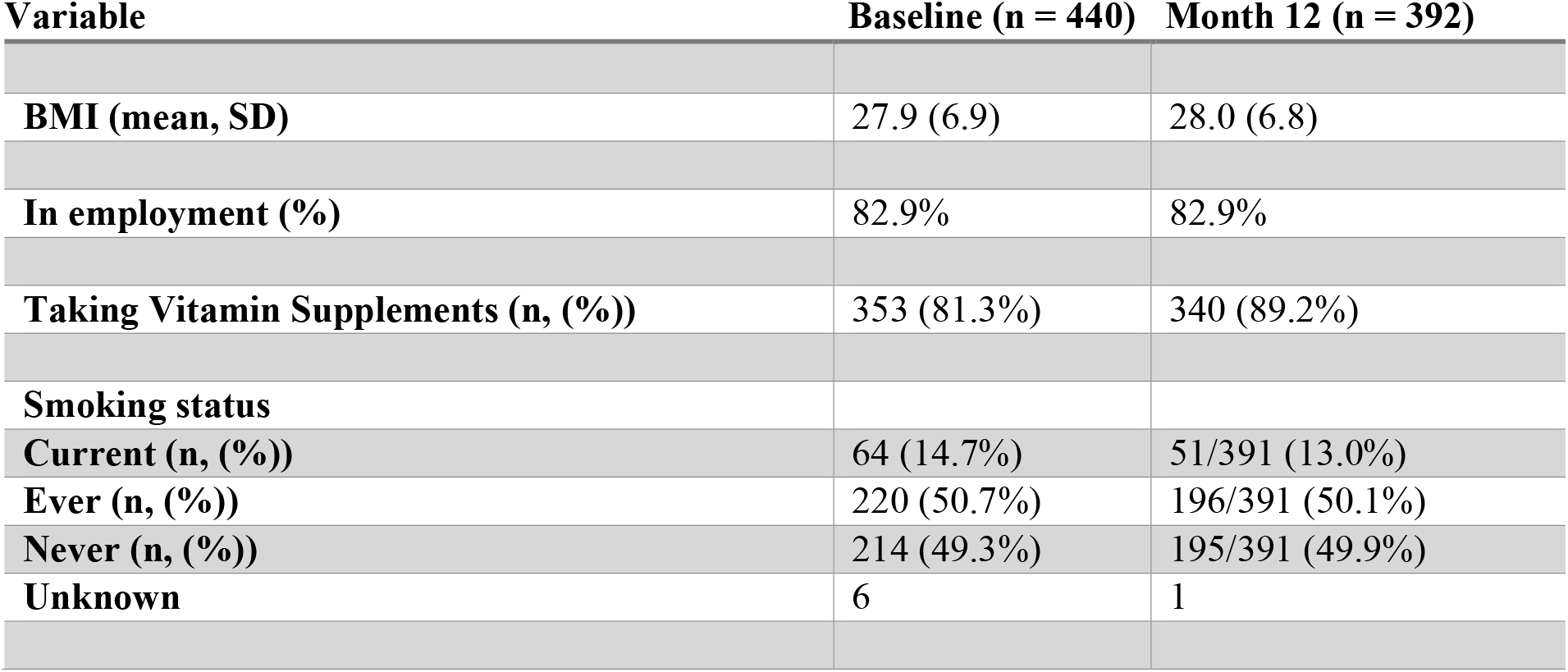
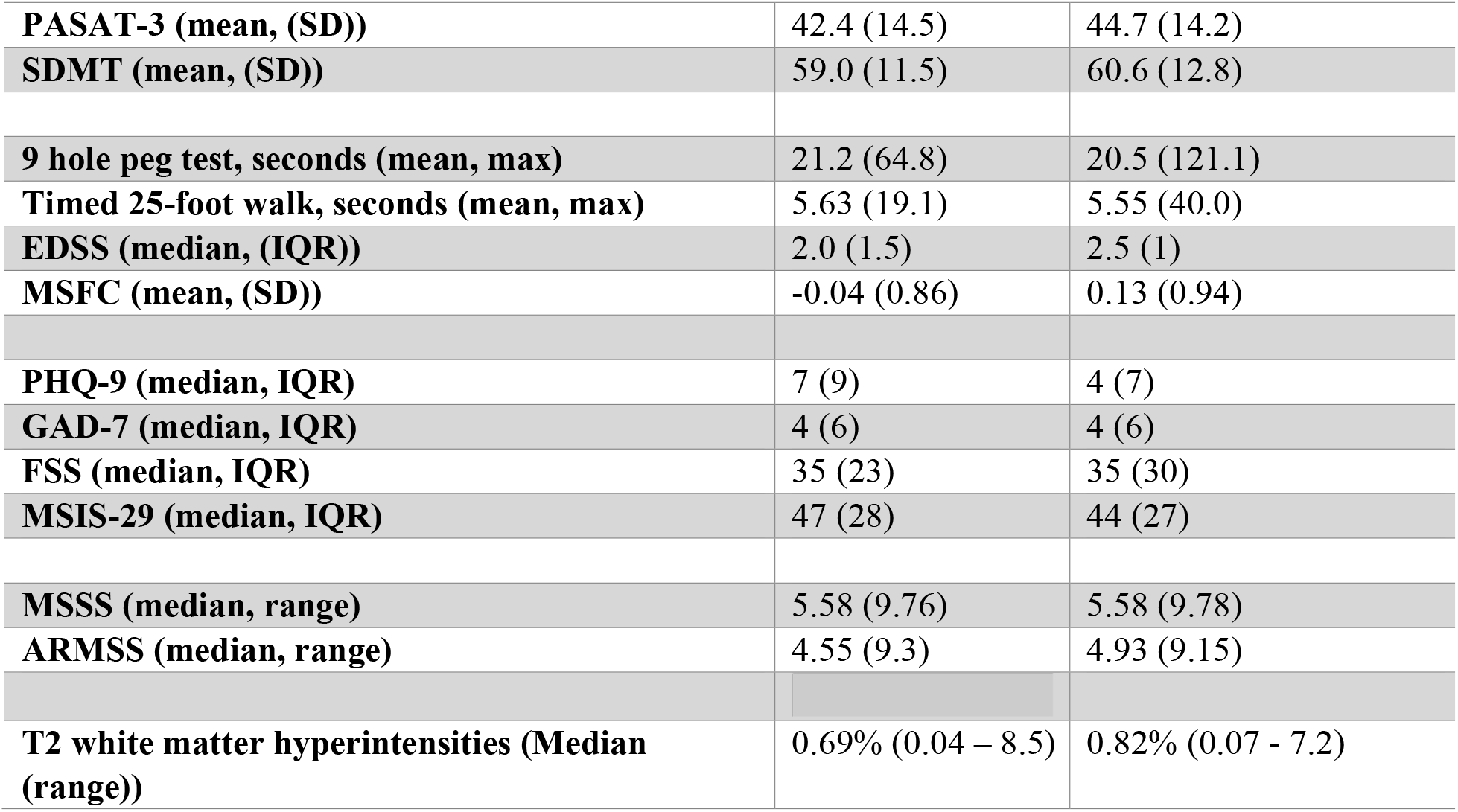
Summary of baseline and month 12 clinical, radiological, and lifestyle measures.

| Variable | Baseline (n = 440) | Month 12 (n = 392) |
| --- | --- | --- |
| **BMI (mean, SD)** | 27.9 (6.9) | 28.0 (6.8) |
| **In employment (%)** | 82.9% | 82.9% |
| **Taking Vitamin Supplements (n, (%))** | 353 (81.3%) | 340 (89.2%) |
| **Smoking status** |  |  |
| **Current (n, (%))** | 64 (14.7%) | 51/391 (13.0%) |
| **Ever (n, (%))** | 220 (50.7%) | 196/391 (50.1%) |
| **Never (n, (%))** | 214 (49.3%) | 195/391 (49.9%) |
| **Unknown** | 6 | 1 |
| **PASAT-3 (mean, (SD))** | 42.4 (14.5) | 44.7 (14.2) |
| **SDMT (mean, (SD))** | 59.0 (11.5) | 60.6 (12.8) |
| **9 hole peg test, seconds (mean, max)** | 21.2 (64.8) | 20.5 (121.1) |
| **Timed 25-foot walk, seconds (mean, max)** | 5.63 (19.1) | 5.55 (40.0) |
| **EDSS (median, (IQR))** | 2.0 (1.5) | 2.5 (1) |
| **MSFC (mean, (SD))** | -0.04 (0.86) | 0.13 (0.94) |
| **PHQ-9 (median, IQR)** | 7 (9) | 4 (7) |
| **GAD-7 (median, IQR)** | 4 (6) | 4 (6) |
| **FSS (median, IQR)** | 35 (23) | 35 (30) |
| **MSIS-29 (median, IQR)** | 47 (28) | 44 (27) |
| **MSSS (median, range)** | 5.58 (9.76) | 5.58 (9.78) |
| **ARMSS (median, range)** | 4.55 (9.3) | 4.93 (9.15) |
| **T2 white matter hyperintensities (Median (range))** | 0.69% (0.04 – 8.5) | 0.82% (0.07 - 7.2) |

**Table 3:**
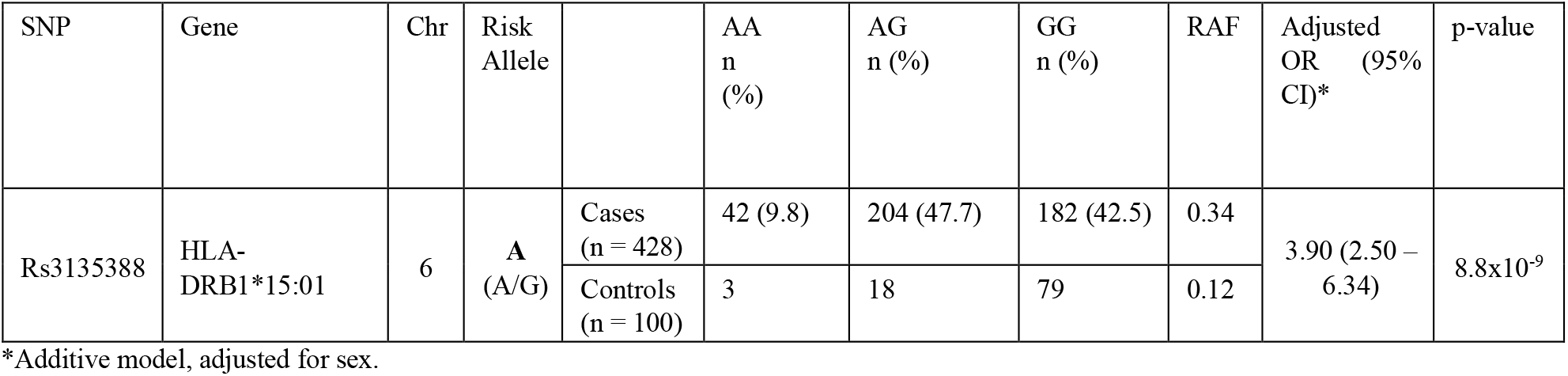
Frequency of HLA-DRB1*15:01 in FutureMS Cases and Controls. OR assume an additive logistic regression model. P-values adjusted for multiple testing by Bonferroni method. RAF = Risk Allele Frequency. SNPs named relative to forward strand Gb37.

However, as shown in Figure 10, despite dominating the contribution to MS risk, the HLA-DRB1*15:01 genotype does not explain much, if any, of the baseline heterogeneity in the age at diagnosis, measured disability severity, or participant reported disease impact in the FutureMS cohort. This underscores that risk genes (HLA-DRB1*15:01) may not necessarily intersect with the gene set that influences disease course. 713,026 SNPs are available for genome-wide analyses in the FutureMS cohort from successful genotyping of 427/428 cases and 100/100 controls for whom PBMCs were available for DNA extraction (see supplemental table and detailed methods). Investigation of the genetic and gene-environment interactions that explain heterogeneity and personal disease trajectories is a focus of ongoing analyses.

**Figure 8.**
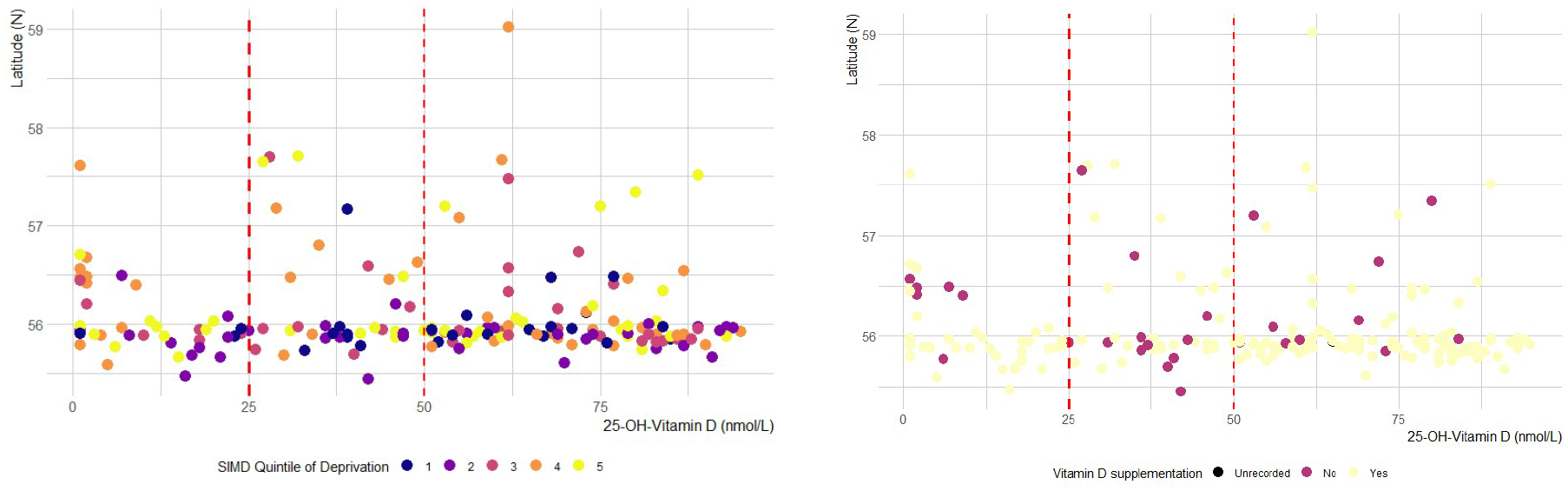
Vitamin D and Latitude of residence show no relationship for the subset (n = 186) of participants with 25-OH-Vitamin D measured at baseline visit (>80% of participants recorded as taking vitamin D supplements). Univariate linear regression shows no relationship with latitude of residence or quintile of socioeconomic deprivation. SIMD quintile of deprivation (1 = most deprived, 5 = least deprived).

**Figure 10:**
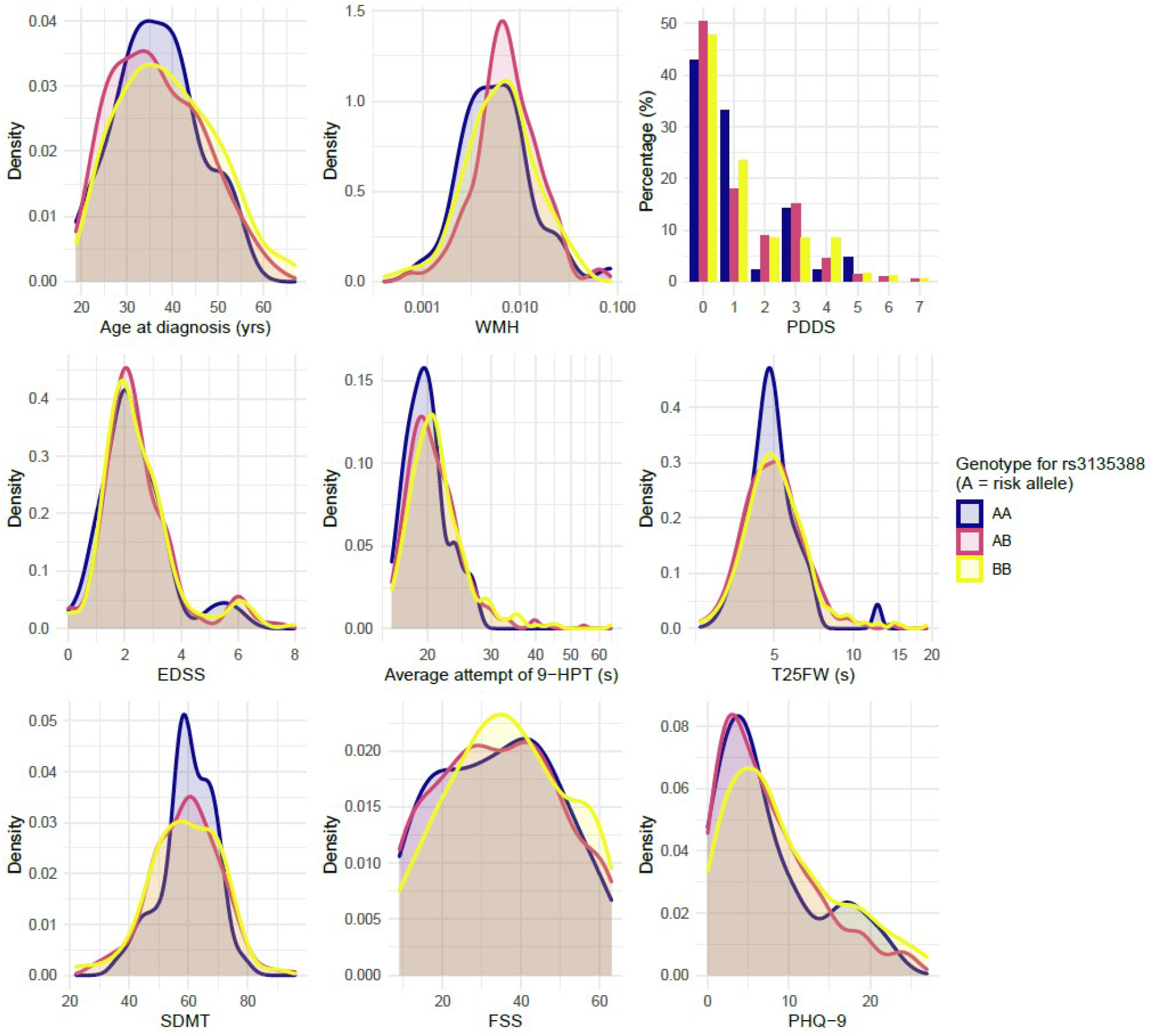
Clinical and radiological measures at baseline visit stratified by HLA-DRB1*15:01 genotype.

## GENOTYPING MATERIALS AND METHODS

Extracted DNA was normalised to 50ng/ul after quantification using Qubit. Samples were genotyped using Infinium HTS chemistry and Infinium Global Screening Array-24 kit. Arrays were scanned on an Illumina iScan system and genotypes were called using GenomeStudio v2.0.3. Genotype calls using GenCall (v6.3.0) with a cut-off specified at 0.15, were then manually reviewed within Genome Studio, using a rigorous multi-step appraisal of cluster fit based on cluster separation score, call frequency, heterozygous excess, heterozygous mean normalized intensity and theta, and minor allele frequency. This was in line with manufacturer published instructions. Further QC was performed using PLINK v1.07 and R v3.5.2.

### Data Management

Participants were identified with a unique non-identifiable study number, which was used to label all paperwork, biological samples and imaging obtained throughout the duration of the study. Questionnaires and clinical data were entered in real time to a FutureMS electronic case report form via an online platform. Data were managed in accordance with the Data Protection Act (DPA 1998), NHS Scotland, and University of Edinburgh policies (supplemental figure).

### Missing Data Handling

Most (395/440) participants in the study recorded entirely complete (100%) baseline records comprising 189 variables in the core clinical dataset. Similar completeness of data was observed for month 12: >99% across all clinical measured and reported variables at both baseline and month 12 follow up. Where missing, source data were carefully inspected for clinical measures/variables and the likely cause for missingness was appraised by a multidisciplinary study team (study nurses and neurologists). Where data were missing at random, multiple imputation with chained equations by predictive mean matching was used to impute baseline measures from across the cohort. Data missing not at random (e.g. due to disability) were left missing where appropriate (e.g. for smoking status) or substituted where appropriate (e.g. when missing timed 25ft walk test due to EDSS > 6 a low z-score was substituted to reflect this disability based on previous cohorts).

### Data Retention

Data acquired in FutureMS may be of potential long-term scientific value. All data collected will therefore be retained for a minimum of 30 years after study completion. Collected data will also be retained after the withdrawal of participants for any reason including loss of capacity. No identifiable data will be shared with third parties, but proposals for collaborative ethically approved research projects utilising these data will be welcomed and proposals considered.

### Statistical analyses

Mixed effects regression models, latent class/transition models, network-based analyses are planned for subsequent investigation of relationships between variables and will be explained in detail elsewhere.

### Ethics

The study is conducted in conformity with the declaration of Helsinki, ICH guidelines for Good Clinical Practice (CPMP/ICH/135/95), and with ethical approval granted from the National Health Service (NHS) South East Scotland Research Ethics Committee (02) (Reference: 15/SS/0233) and approval granted from individual NHS Board Research and Development departments (IRAS Project ID: 169955). Control samples were collected under approval granted from the NHS East of Scotland Research Ethics committee (REC01) as part of the Scottish Regenerative Neurology Tissue Bank Project (Reference: 15/ES/0094).

### Funding

FutureMS has been funded by Precision Medicine Scotland Innovation Centre (PMS_IC), the Rowling Clinic, and Biogen Idec. Ltd. Insurance provided by the Co-Sponsors: NHS Lothian and the University of Edinburgh.

## DISCUSSION, STRENGTHS AND LIMITATIONS

We have designed and recruited a large cohort of persons with RRMS across Scotland. The prospective nature of FutureMS enables longitudinal assessment of clinical, imaging, genomic and fluid biomarkers in all participants prior to and during disease modifying treatments. As the number of available treatment options increases, so too must our understanding of the heterogeneity of disease course for persons living with RRMS. Substantial effort has been made to ensure that the study has recruited a geographically, socioeconomically, and clinically as representative a cohort as possible. Results presented here give us confidence that this has been achieved such that findings from this study may generalize to the clinic and real world.

Scotland has long been recognized as having a high incidence of multiple sclerosis, for reasons that remain unknown despite long-running speculation^7^. The Scottish northern isles for many decades have been recognized as particularly burdened^30^. Our early exploration of genetic results confirms expected findings of an excess of HLA-DRB1*15:01 (OR 3.90, 95% CI: 2.50 – 6.34) in the Scottish MS population. This provides a useful prevalence benchmark by which this (and other) genetic loci can be assessed and compared to other MS populations. The risk allele frequency (RAF) in the FutureMS cases of 0.34 is high by previously published standards^31–34^, but not extremely so with numerous historical case control studies reporting higher frequencies of this gene^35^. The frequency in controls (0.12), is similarly high, but not excessively so. Taken together, the excess frequency in cases underscores the highly probable importance of this gene’s contribution to MS risk in Scotland (as elsewhere), but leaves room for other genetic or environmental factors to explain why Scotland has a high rate of MS. Substantial further exploration is required and intended to address this issue.

Recent and historical studies have noted regional variation in the distribution of this burden of MS across Scotland^4,36–38^, consistent with findings in many other countries where regional analyses have been performed^39,40^. A strength of this study is that in being geographically representative of the national population it may be well positioned to investigate genetic and environmental hypotheses for this spatial heterogeneity in disease burden.

The exploratory analyses presented here demonstrate that our cohort can be considered nationally representative. That said, we suggest caution generalizing any findings from this population to individuals who fall outside of he remit of our study. For example, those who experience such aggressive disease at onset that DMT is initiated emergently (as these people would not have been eligible for recruitment had they been commenced on DMTs emergently at the time of diagnosis), or to those diagnosed at extremes of the age distribution (particularly <18). Similarly, caution may be necessary if attempting to generalize to populations with more heterogenous recent ancestry and to those whose initial presentation is with progressive disease.

Multiple sclerosis is a clinically heterogenous disease, presenting with variable symptoms affecting different parts of the central nervous system, which may be interspersed by prolonged periods without overt disease. This heterogeneity makes the diagnosis challenging and often delayed. Variability between clinicians can compound this heterogeneity. Although we used six months as a proxy for ‘newly diagnosed’ this does not necessarily equate to ‘early’ disease from a pathophysiological view, and this is an important limitation of our study. This is shown by the time taken from first symptom to diagnosis ranging from a single day to 33.5 years in the FutureMS cohort. It is perhaps inevitable, therefore that biological markers taken at baseline research will often not be reflect true disease initiation. However, a strength of this study is that participants were enrolled as early as possible after diagnosis. Whilst date of disease diagnosis will not be equivalent to date of disease onset, it is a best practical compromise.

Our early exploration of the association with disability severity and demographic and lifestyle factors highlighted an obvious difference, observable at baseline, in measures of physical disability between current and non-smokers. Importantly, the proportion of current smokers barely changed over the first year of the study, despite the wealth of evidence of the risk of smoking worsening disease control. This brings into focus the need to counsel all persons newly-diagnosed with MS who smoke, as early as possible, and to provide information on the benefits of cessation and the MS-specific harm of smoking, including passive smoking (which we didn’t capture). It is likely that for some pwMS, particularly those for whom smoking is compounded with genetic pre-disposition, the effect of stopping smoking may be very substantial.

Fortunately, depression as measured by PHQ-9 is one of the clinical measures that improves most in the first year following diagnosis. However, we noted a high burden of high PHQ-9 scores at baseline, and particularly in persons diagnosed with MS who are also unemployed. Numerous possible explanations for the relationship between MS-depression and employment are possible, and further work will be necessary and is intended to delineate the causal structure of this relationship in order to guide effective treatment. However, these findings underscore the importance of considering the burden of mental health conditions in MS. It is reassuring that these scores improve on average in the early phase of the condition, and this may be of reassurance to some patients, and may encourage mental health treatment where mood is not improving.

In conclusion, we anticipate that long-term follow up of the FutureMS cohort will lead to the development and implication of clinical tools for predicating future disability in patients with multiple sclerosis.

## Data Availability

Data requests from a third party will be handled on a case by case basis by the steering committee and data custodians.

## ACKNOWLEDGEMENTS

This study is indebted to the FutureMS participants. We would like to thank non-author contributors of the FutureMS Consortium and clinical collaborators within neurology departments across NHS Scotland. With thanks to FutureMS, hosted by Precision Medicine Scotland Innovation Centre [PMSIC] and funded by a grant from the Scottish Funding Council to Precision Medicine Scotland Innovation Centre (PMS IC) and Biogen Idec Ltd Insurance. PK is supported by ECAT/Wellcome fellowship. Many thanks to Olivia Fleming for critical review of the manuscript.

## SUPPLEMENTARY FIGURES AND TABLES

**Figure S1.**
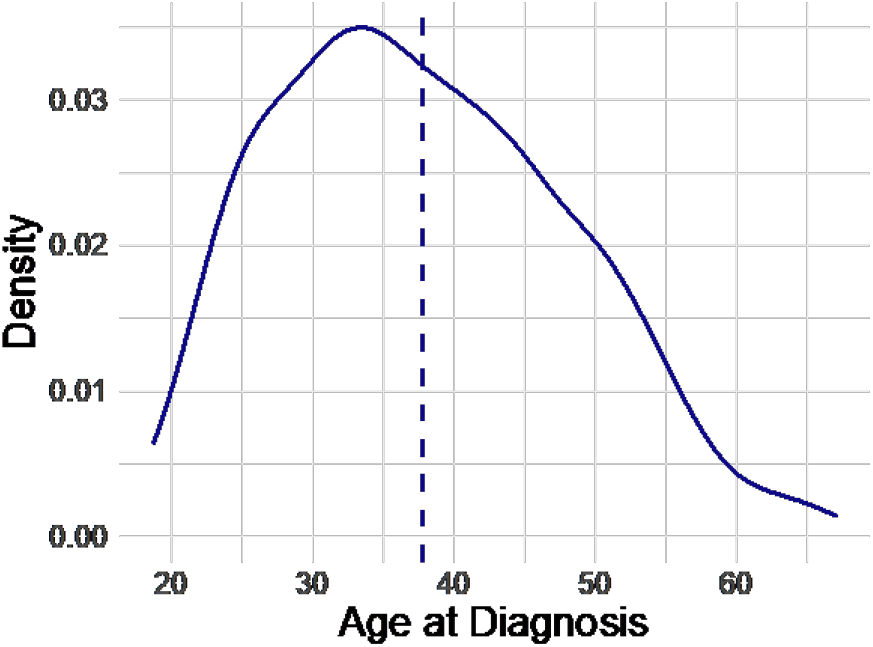
Demonstrating truncation at extremes of age (particularly <18) due to inclusion criteria. Will mention in limitations.

**Table S1.** Summary of study visit.

| ASSESSMENT | Baseline | Year 1 | Year 5 (in planning) |
| --- | --- | --- | --- |
| Informed Consent | **X** |  | **X** |
| Demographics | **X** |  |  |
| Medical History | **X** | **X** | **X** |
| Social History | **X** | **X** | **X** |
| Family History | **X** | **X** | **X** |
| Migration History |  |  | **X** |
| Relapse/Progression History | **X** | **X** | **X** |
| Medication History | **X** | **X** | **X** |
| Structured Neurological Examination | **X** | **X** | **X** |
| BMI, Height, Weight, BP | **X** | **X** | **X** |
| Measures of Physical Disability (EDSS, T25FTWT, 9HPT) | **X** | **X** | **X** |
| Cognitive tests (PASAT-3, SDMT) | **X** | **X** | **X** |
| PHQ-9 and GAD-7 | **X** | **X** | **X** |
| MSIS-29 | **X** | **X** | **X** |
| Fatigue Severity Score | **X** | **X** | **X** |
| Patient Derived Disease Severity Score | **X** | **X** | **X** |
| Visual Acuity | **X** | **X** | **X** |
| Standard MRI Protocol | **X** | **X** | **X** |
| Disease Modifying Therapy History | **N/A** | **X** | **X** |
| Lymphocyte subsets stored (CD4, CD8, CD20, CD??) | **X** |  |  |
| DNA Genotyping | **X** |  |  |
| Advanced MRI Imaging (SS3) | **X** | **X** | **X** |
| Optical Coherence Tomography (SS4) | X | X | X |

**Figure S2.**
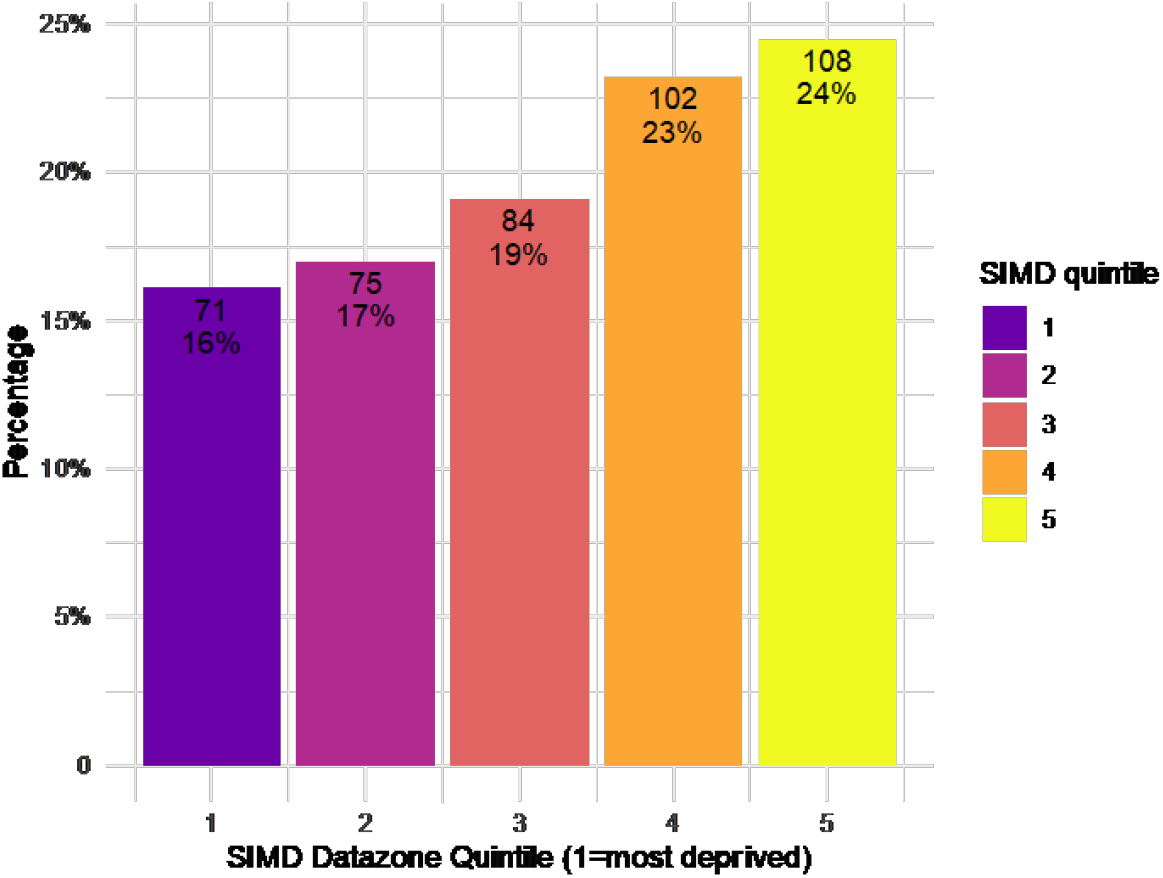
SIMD quintiles.

**Figure S3:**
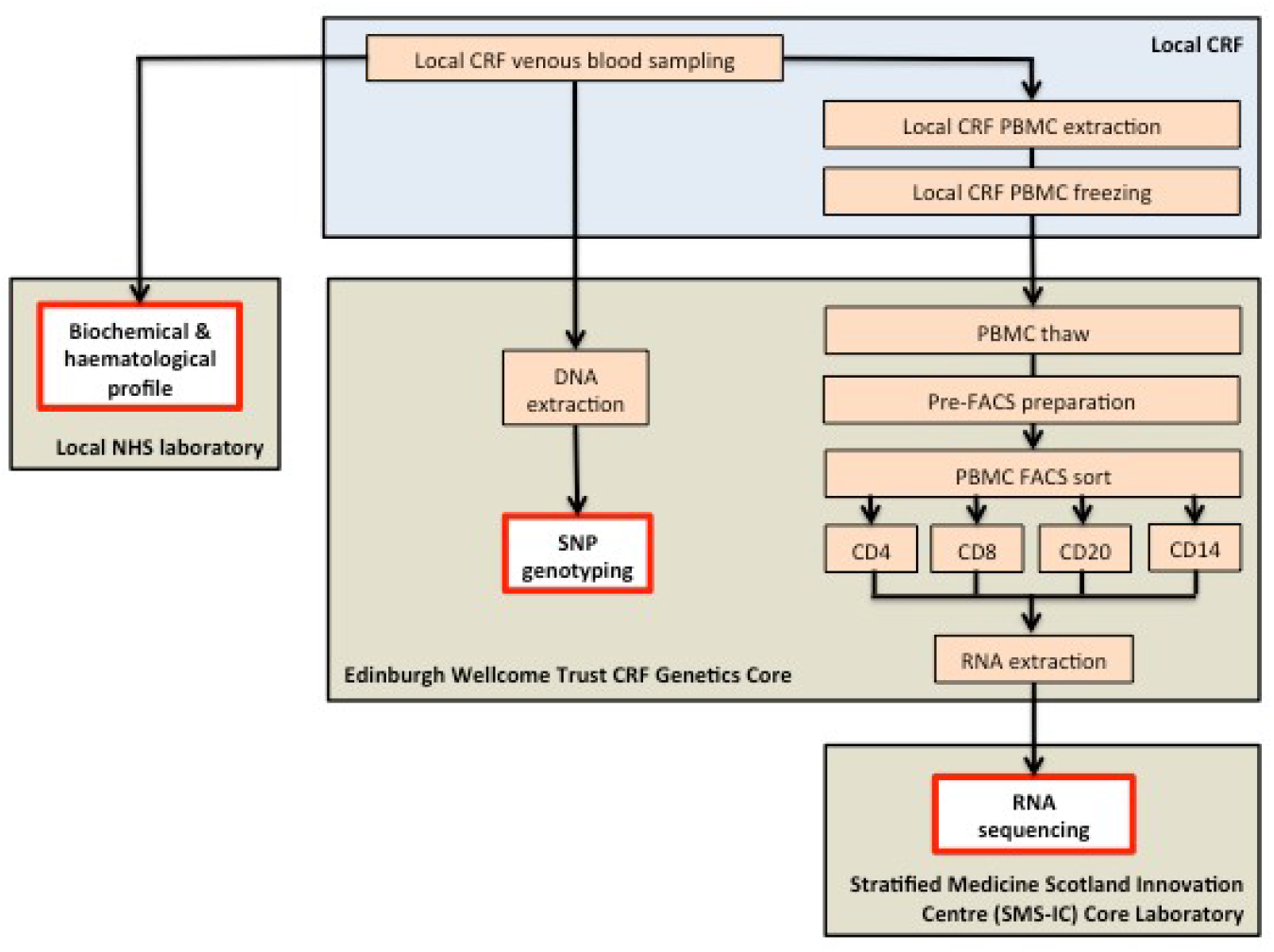
Schematic explaining laboratory pathway for tissue collection for biomarker processing.

**Supplementary table 2:** Summary of numbers of FutureMS study subjects and controls with peripheral blood mononuclear cells available for analysis.

|  | **With PBMCs** | **FACs sorted** | **Total** |
| --- | --- | --- | --- |
| Edinburgh (01) | 184 | 184 | 185 |
| Glasgow (02) | 163 | 137 | 165 |
| Dundee (03) | 46 | 46 | 46 |
| Aberdeen (04) | 35 | 32 | 35 |
| Inverness (05) | 8 | 6 | 8 |
| Controls | 100 | 78 | 103 |
| **Totals (patients only)** | **436** | **405** | **439** |
| **Totals** | **536** | **483** | **542** |

**Figure S4.**
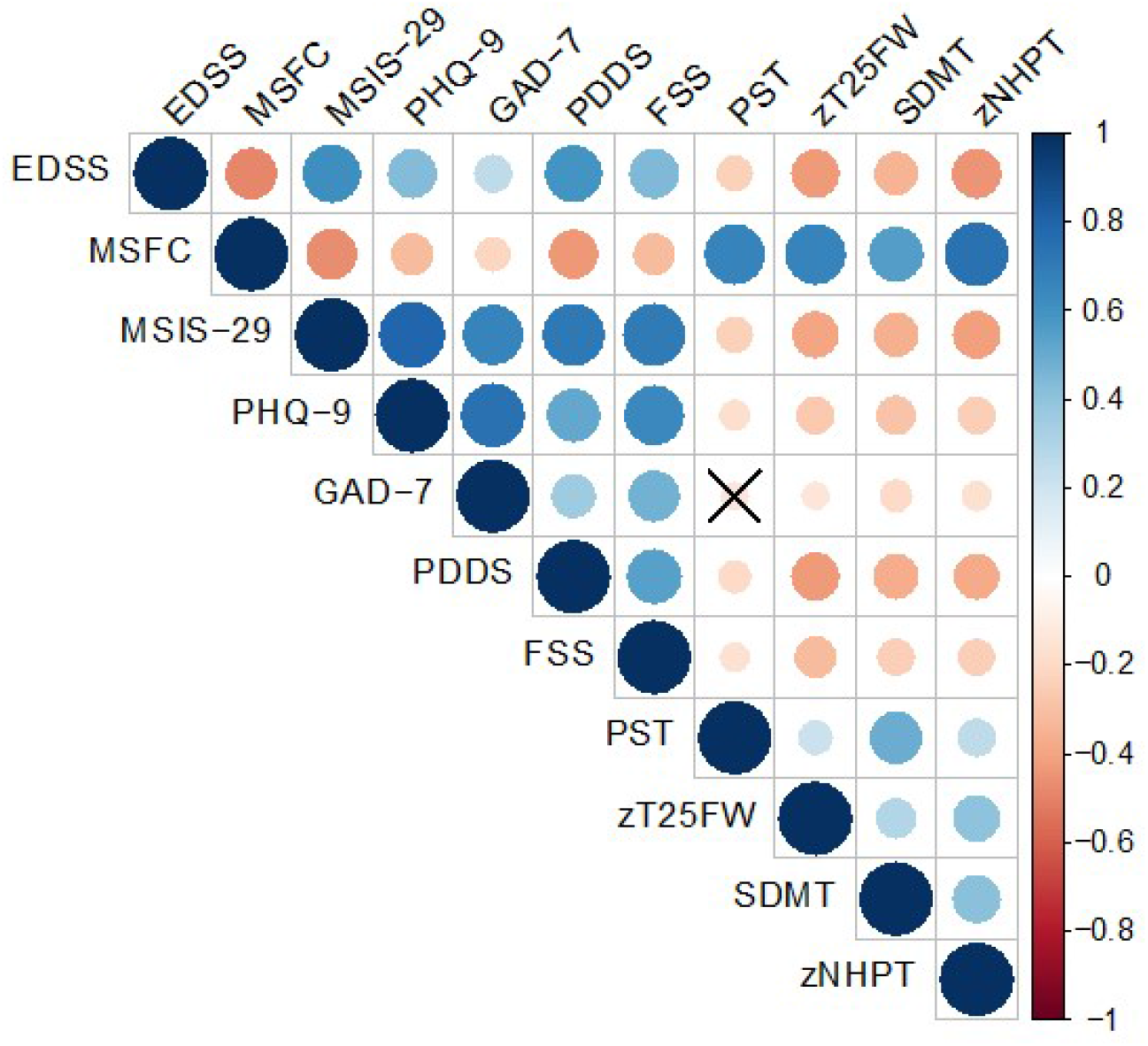
Correllelogram of clinical measures at baseline visit.

**Supplementary Figure 5.**
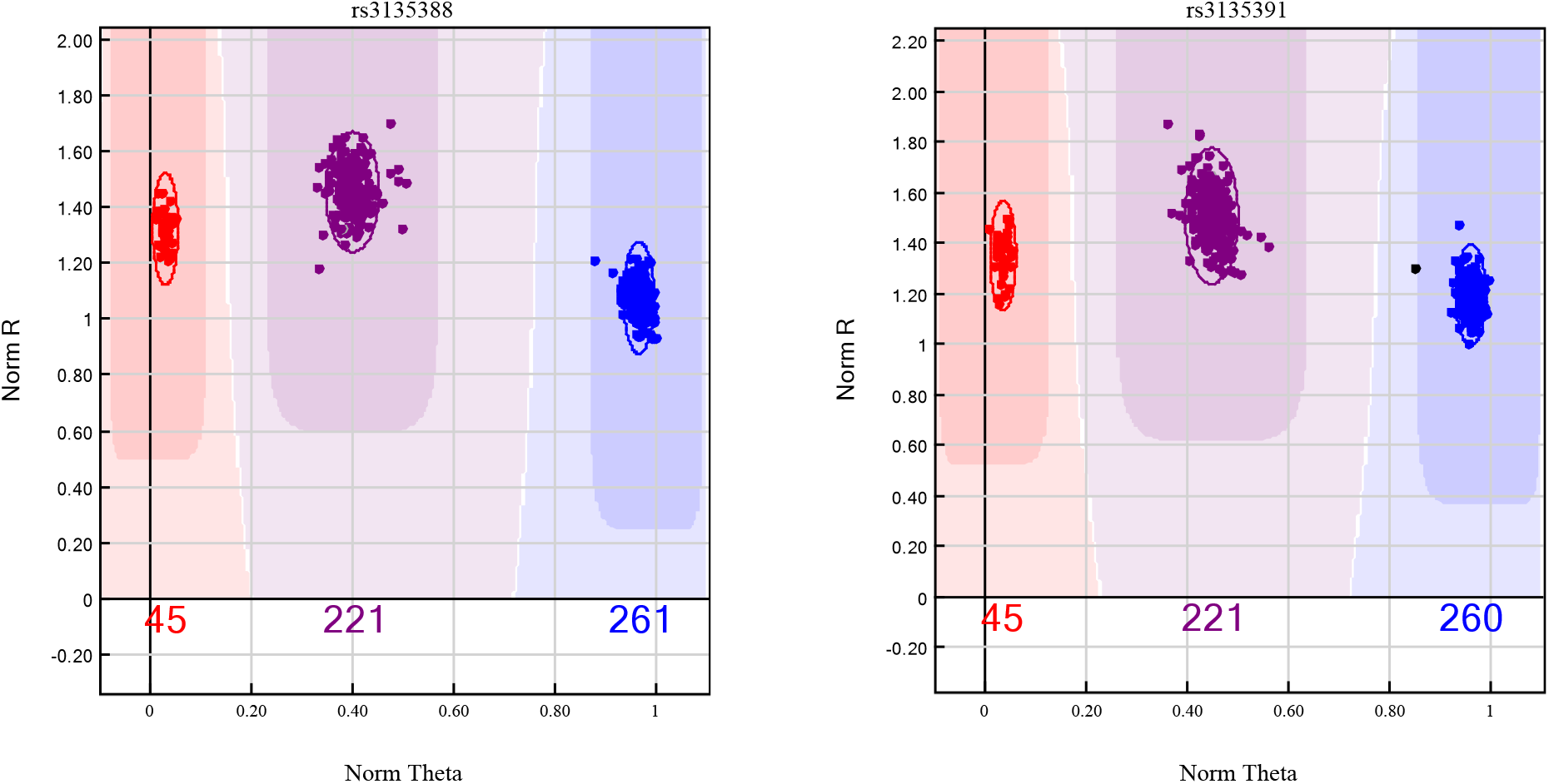
Demonstration of two SNPs in near perfect linkage disequilibrium from the FutureMS cohort demonstrating clear separation of clusters resulting in high confidence of genotype calls. These two SNPs mark HLA-DRB1*15:01, which contributes the largest single gene effect for multiple sclerosis.

**Supplementary table 3.** Proportion of PBMCs expressing markers for monocytes, B cells, helper and cytotoxic t cells in the peripheral blood mononuclear cells donated by patients to thestudy.

|  | Parent/Grandparent | % of Parent |  |  |  | % of Grandparent |  |  |  |
| --- | --- | --- | --- | --- | --- | --- | --- | --- | --- |
|  |  | Mean | SD | LQ10 | UQ10 | Mean | SD | LQ10 | UQ10 |
| Single cells | Events | 56.05 | 15.98 | 35.60 | 75.00 |  |  |  |  |
| Viable (7AAD -ve) | Single cells | 91.77 | 10.11 | 97.80 | 84.93. |  |  |  |  |
| CD14+ Monocytes | CD3-/viable cells | 22.03 | 11.67 | 34.30 | 10.40 |  |  |  |  |
| CD19+ B cells | CD3-/viable cells | 8.11 | 3.69 | 4.08 | 12.9 |  |  |  |  |
| CD4+ T cells | CD3+/viable cells | 69.44 | 11.67 | 81.47 | 57.00 | 38.97 | 10.48 | 51.27 | 26.56 |
| CD8+ T cells | CD3+/viable cells | 20.27 | 8.31 | 30.37 | 11.22 | 20.27 | 8.31 | 30.37 | 11.22 |

**Supplementary figure 6.**
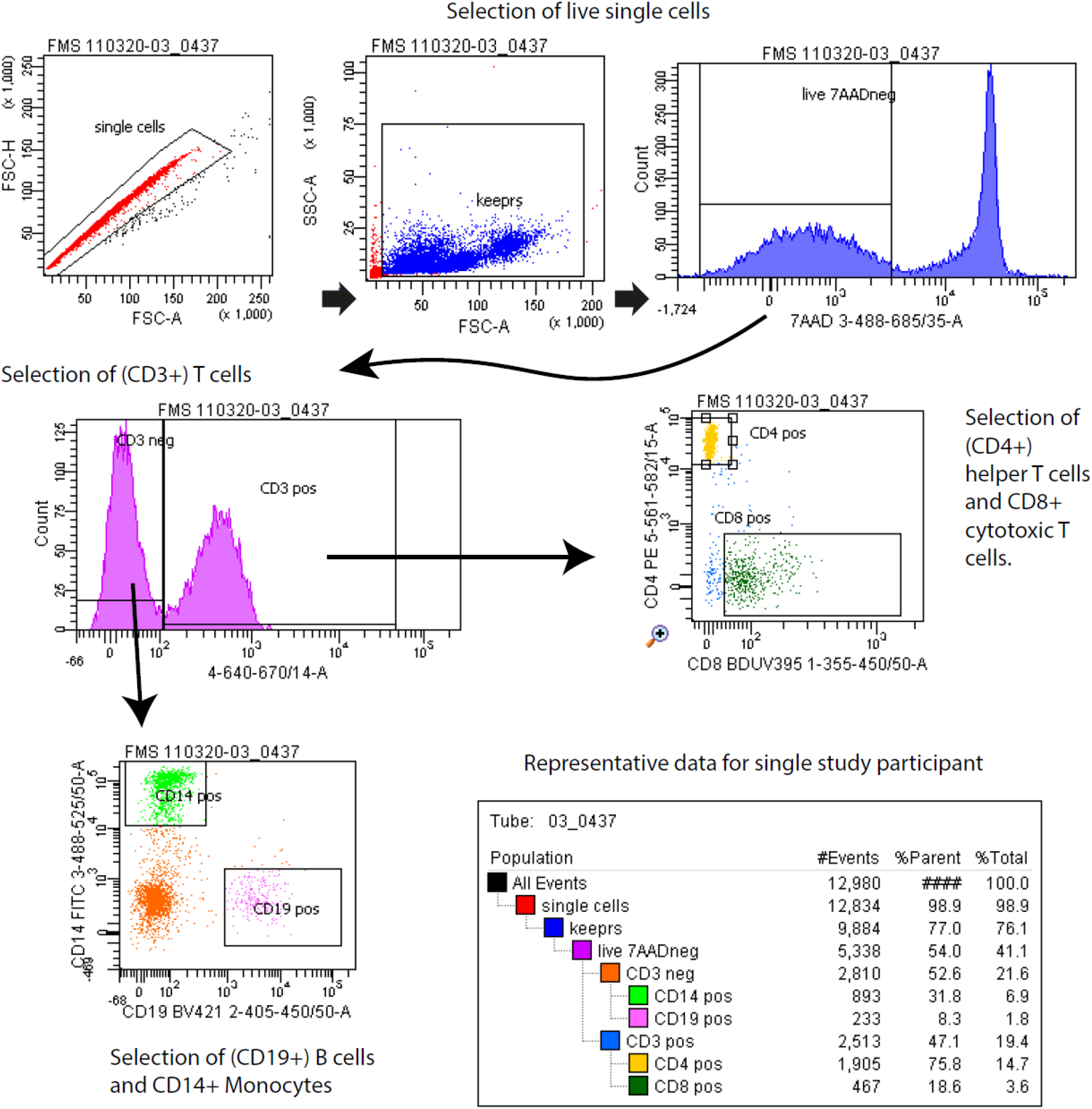
Example of the fluorescence-activated cell sorting undertaken for a representative member of the FMS cohort. CD3 – T cell marker. CD4+ helper T cells, CD8+ cytotoxic T cells. CD19+ B cell marker and CD14+ monocyte marker. 7AAD – 7-Aminoactinomycin D uptake associated with loss of cell viability.

